# Lower Omega-3 Status Associated with Higher Erythrocyte Distribution Width and Neutrophil-Lymphocyte Ratio in UK Biobank Cohort

**DOI:** 10.1101/2022.12.09.22283290

**Authors:** Michael I. McBurney, Nathan L. Tintle, William S. Harris

## Abstract

High red blood distribution width (RDW) is associated with decreased red blood cell deformability, and high neutrophil-lymphocyte ratio (NLR) is a biomarker of systemic inflammation and innate-adaptive immune system imbalance. Both RDW and NLR are predictors of chronic disease risk and mortality. Omega-3 index (O3I) values have previously been shown to be inversely associated with RDW and NLR levels. Our objective was to determine if total plasma long chain omega-3 fatty acids (Omega3%) measured in the UK Biobank cohort were associated with RDW and NLR values. RDW- and NLR-relationships with Omega3% were characterized in 109,191 adults (58.4% female). RDW- and NLR-Omega3% relationships were inversely associated with Omega3% (both p<0.0001). These cross-sectional associations confirm previous findings that increasing RDW and NLR values are associated with low O3I. The hypothesis that RDW and/or NLR values can be reduced in individuals with less-than optimal long chain omega 3 values need to be tested in randomized controlled intervention trials using EPA and/or DHA.

## Introduction

A complete blood count is a routine component of medical exams that provides comprehensive hematologic information. Using standardized reference ranges, healthy individuals can be distinguished from those with nutrient deficiencies, infections, tissue damage, and inflammation. In addition to basic information provided by hemoglobin, hematocrit, and platelet and white blood cell counts, the red blood cell distribution width (RDW) is routinely used to diagnose anemias [1] and may be useful to assess overall wellness [2]. Another metric, the neutrophil-to-lymphocyte ratio (NLR), a measure of innate-adaptive immune balance, is used to monitor inflammation and immunity [3–6]. Risk of non-communicable disease and mortality has been associated with elevated RDW (**Table 1**) and NLR values (**Table 2**).

**Table 1.** Odds Ratio (OR) or Hazards Ratio (HR) per 0.01 unit increase in red blood cell distribution width (RDW) with respect to clinical outcome.

| Outcome | Population | Study | n | Quantile of RDW |  |  |  |  | OR or HR<br>(95% CI) | p-value | Citation |
| --- | --- | --- | --- | --- | --- | --- | --- | --- | --- | --- | --- |
|  |  |  |  | 1 | 2 | 3 | 4 | 5 |  |  |  |
| Frailty | Hospitalized men, ≥60y | Comprehensive tertiary hospital in mainland China | 334 | ≤12.6% | 12.6-13.1% | 13.1-13.7% | >13.7% |  | OR=1.14<br>(1.05-1.24) | 0.002 | [32] |
| Frailty | Hospitalized women, ≥60y | Comprehensive tertiary hospital in mainland China | 285 | ≤12.6% | 12.6-13.1% | 13.1-13.7% | >13.7% |  | OR=1.27<br>(1.16-1.38) | <0.001 | [32] |
| Incident atrial fibrillation (AF) | Adults ≥25y | inhabitants of Tromso, Norway | 26,111 | <12.3% | 12.4-12.7% | 12.8-13.2% | ≥13.3% |  | HR=1.08<br>(1.03-1.12) | Not reported | [33] |
| Venous thromboembolism | Adults with AF ≥25y | inhabitants of Tromso, Norway | 1,812 | <12.7% | 12.8-13.3% | >13.3% |  |  | HR=1.09<br>(0.83-1.41) | Not reported | [33] |
| Ischemic stroke | Adults with AF ≥25y | inhabitants of Tromso, Norway | 1,812 | <12.7% | 12.8-13.3% | >13.3% |  |  | HR=1.18<br>(1.03-1.34) | Not reported | [33] |
| In-hospital mortality | Patients without anemia who underwent percutaneous coronary intervention (PCI) | Affiliated hospital of Zhengzhou University, China | 2,169 | <12.27% | 12.27-≤13.0% | >13-<13.5% | ≥13.5% |  | HR=1.4<br>(1.23-1.59) | 0.000 | [34] |
| All-cause mortality | Patients with stable coronary artery disease undergoing stent implantation | Silesian Centre for Heart Diseases, Poland | 2,033 | <13.1% | 13.1-<13.6% | 13.6-<14.1% | ≥14.1% |  | HR=1.23<br>(1.13-1.35) | <0.0001 | [35] |
| All deaths, men | Adults ≥20y without diabetes or CV | NHANES 1988-1994, USA | 7,201 | ≤12.4% | >12.4-≤12.9% | >12.9-≤13.5% | >13.5% |  | HR=1.21<br>(1.18-1.25) | <0.001 | [36] |
| All deaths, women | Adults ≥20y without diabetes or previous CV | NHANES 1988-1994, USA | 8,259 | ≤12.4% | >12.4-≤12.9% | >12.9-≤13.5% | >13.5% |  | HR=1.19<br>(1.14-1.24) | <0.001 | [36] |
| All CV deaths, men | Adults ≥20y without diabetes or previous CV | NHANES 1988-1994, USA | 7,201 | ≤12.4% | >12.4-≤12.9% | >12.9-≤13.5% | >13.5% |  | HR=1.19<br>(1.14-1.25) | <0.001 | [36] |
| All CV deaths, women | Adults ≥20y without diabetes or previous CV | NHANES 1988-1994, USA | 8,259 | ≤12.4% | >12.4-≤12.9% | >12.9-≤13.5% | >13.5% |  | HR=1.19<br>(1.12-1.26) | <0.001 | [36] |
| Coronary heart disease (CHD) deaths, men | Adults ≥20y without diabetes or previous CV | NHANES 1988-1994, USA | 7,201 | ≤12.4% | >12.4-≤12.9% | >12.9-≤13.5% | >13.5% |  | HR=1.20 (1.12-1.28) | <0.001 | [36] |
| CHD deaths, women | Adults ≥20y without diabetes or previous CV | NHANES 1988-1994, USA | 8,259 | ≤12.4% | >12.4-≤12.9% | >12.9-≤13.5% | >13.5% |  | HR=1.20 (1.11-1.29) | <0.001 | [36] |
| Peripheral artery disease | Adults ≥40y | NHANES 1999-2004, USA | 6,950 | <12.2% | 12.3-12.5% | 12.6-13.1% | >13.1% |  | HR=1.23 (1.14-1.33) | <0.001 | [37] |
| All-cause mortality | Adults ≥40y | NHANES 1988-1994, USA | 15,852 | <12.35% | 12.5-12.65% | 12.8-13.15% | 13.2-13.75% | ≥13.80% | HR=1.27 (1.21-1.33) | <0.001 | [38] |
| CV mortality | Adults ≥40y | NHANES 1988-1994, USA | 15,852 | <12.35% | 12.5-12.65% | 12.8-13.15% | 13.2-13.75% | ≥13.80% | HR=1.26 (1.19-1.33) | <0.001 | [38] |
| Cancer mortality | Adults ≥40y | NHANES 1988-1994, USA | 15,582 | <12.35% | 12.5-12.65% | 12.8-13.15% | 13.2-13.75% | ≥13.80% | HR=1.30 (1.22-1.39) | <0.001 | [38] |
| Chronic lower respiratory tract disease mortality | Adults ≥40y | NHANES 1988-1994, USA | 15,582 | <12.35% | 12.5-12.65% | 12.8-13.15% | 13.2-13.75% | ≥13.80% | HR=1.36 (1.29-1.51) | 0.03 | [38] |
| All-cause mortality | Adults ≥45y | NHANES 1988-1994, USA | 8,175 | <12.55% | 12.55-12.95% | 13.0-13.4% | 13.45-14.05% | >14.05% | HR=1.22 (1.15-1.30) | <0.001 | [39] |

**Table 2.**
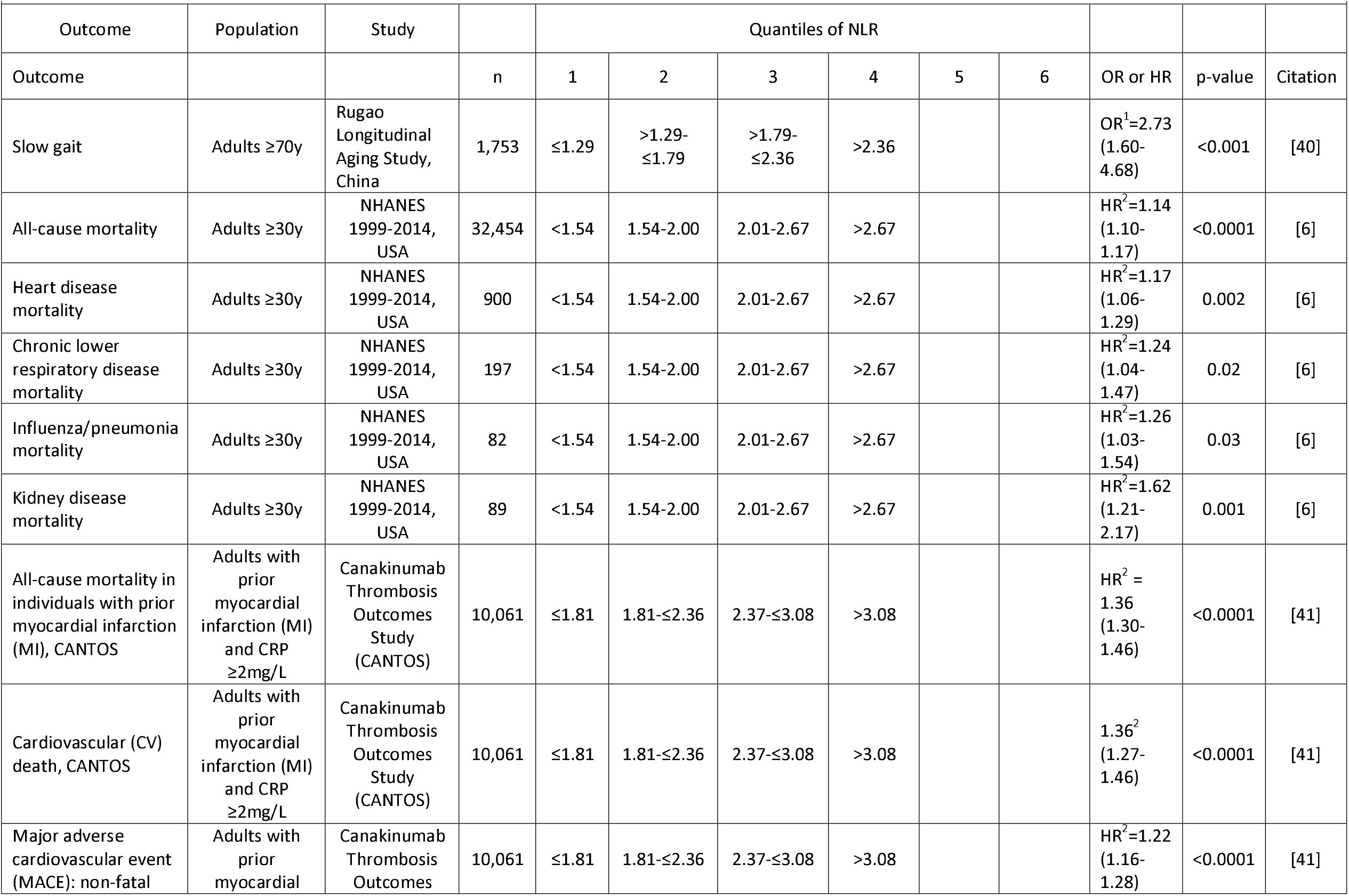
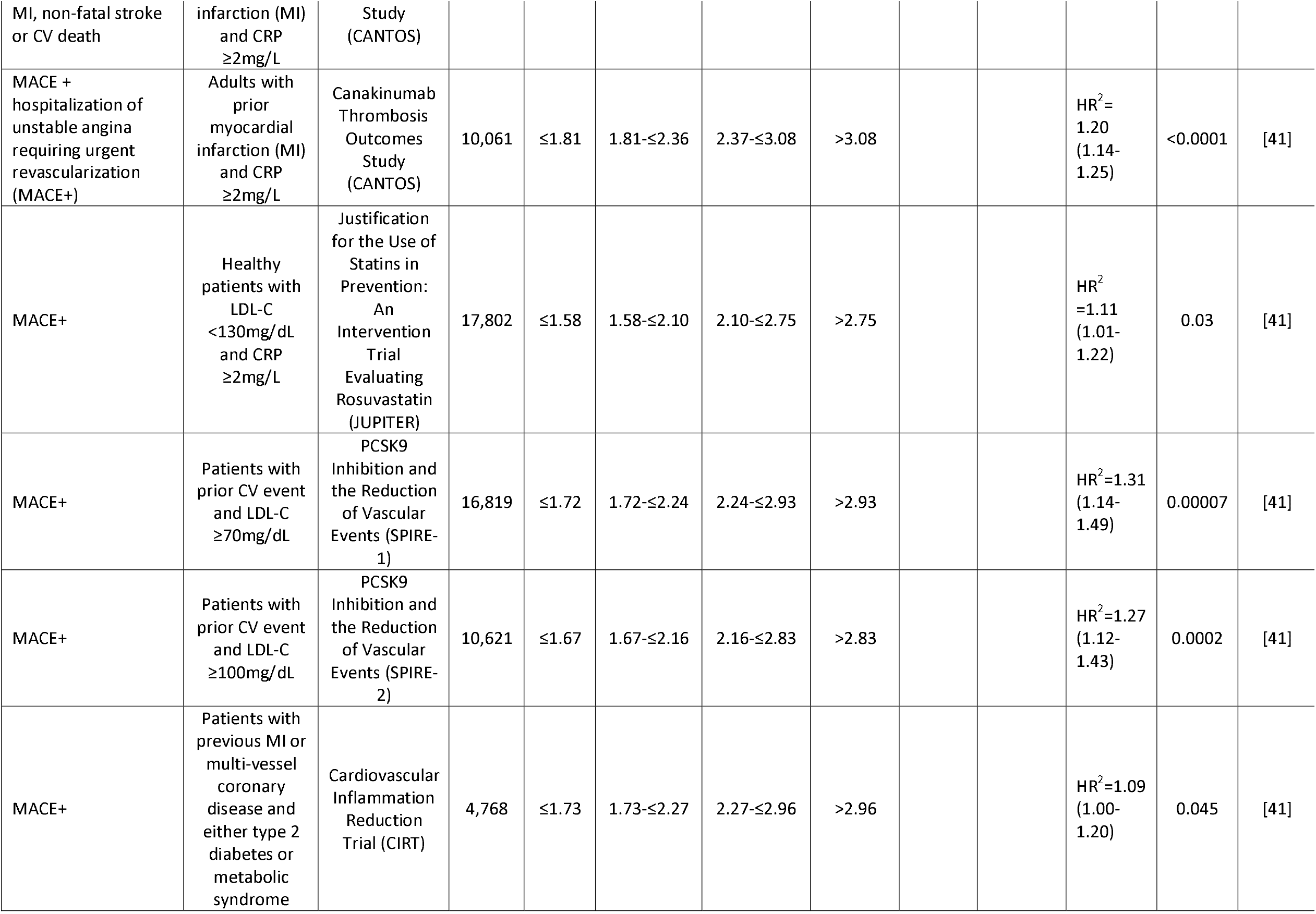
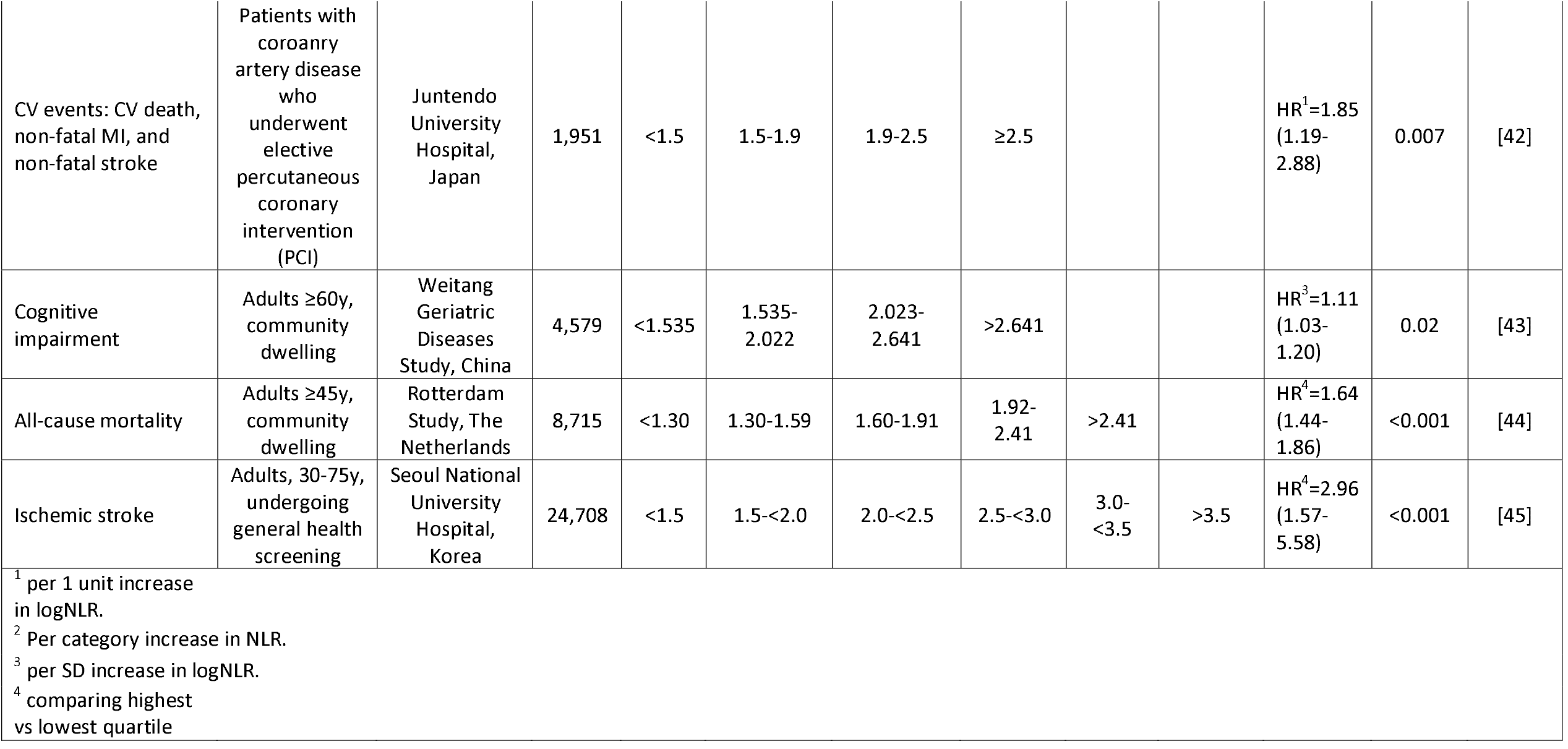
Odds Ratio (OR) or Hazards Ratio (HR) per increase in neutrophil-lymphocyte ratio (NLR) with respect to clinical outcome.

The long-chain omega-3 fatty acid (Omega3%) composition of all cell membranes (including blood cells) affects physicochemical properties such as fluidity, membrane protein function, signaling cascades, overall cell function and ultimately, host physiology [7–9]. The EPA DHA content of RBC membranes, i.e., the Omega-3 Index (O3I) [10], reflects long-term dietary omega-3 intake [11] and the EPA+DHA composition of major organs [12]. Optimal O3I levels for cardiovascular health appear to be >8% [13]. O3I is predictive of risk of total mortality in individuals without prevalent cardiovascular disease [14] and associated with mortality risk before and after adjustment for individual n-6 fatty acids, whether in aggregate, by carbon-chain groups, or individually [15]. A harmonized, de novo analysis of over 42,000 individuals prospectively followed for over 16 years, with over 15,000 deaths observed, found risk for total mortality was 15-18% lower in the highest vs lowest quintile for long chain omega-3 fatty acids and similar relationships were observed for death from cardiovascular disease and cancer [16]. Increased dietary long chain omega-3 fatty acid intake increases red blood cell deformability [17–20], and frequent fish consumption is associated with lower NLR [21].

In previous studies using a clinical US laboratory database, a low O3I was inversely associated with RDW [22] and NLR [23]. The present study aims to confirm reported RDW and NLR relationships with Omega3% in a large and well-characterized cohort from the UK.

## Methods

### Sample

The UK Biobank is a prospective, population-based cohort of 502,639 individuals, aged 40-69y, recruited between April 2007 and December 2010 [24,25]. Total long chain omega-3 fatty acids (Omega3%) and DHA (DHA%) were measured in plasma as a percent of total fatty acids by nuclear magnetic resonance (NMR, Nightingale Health Plc, Helsinki, Finland) [26] at enrollment. Within the cohort, 117,351 adults ≥18y had data available for Omega3% and DHA%, and 109,191 also had complete data on RDW, neutrophil count, lymphocyte count, NLR, age, sex, body mass index (BMI), high-sensitivity C-reactive protein (CRP), and hemoglobin (Hb) (**Figure 1**). Since O3I was not measured in the UK Biobank, the O3I was calculated from Omega3% using the method of Schuchardt et al [27]. UK Biobank has ethical approval (Ref. 11/NW/0382) from the Northwest Multi-centre Research Ethics Committee as a Research Tissue Bank (RTB). All participants gave electronic signed informed consent.

**Figure 1.**
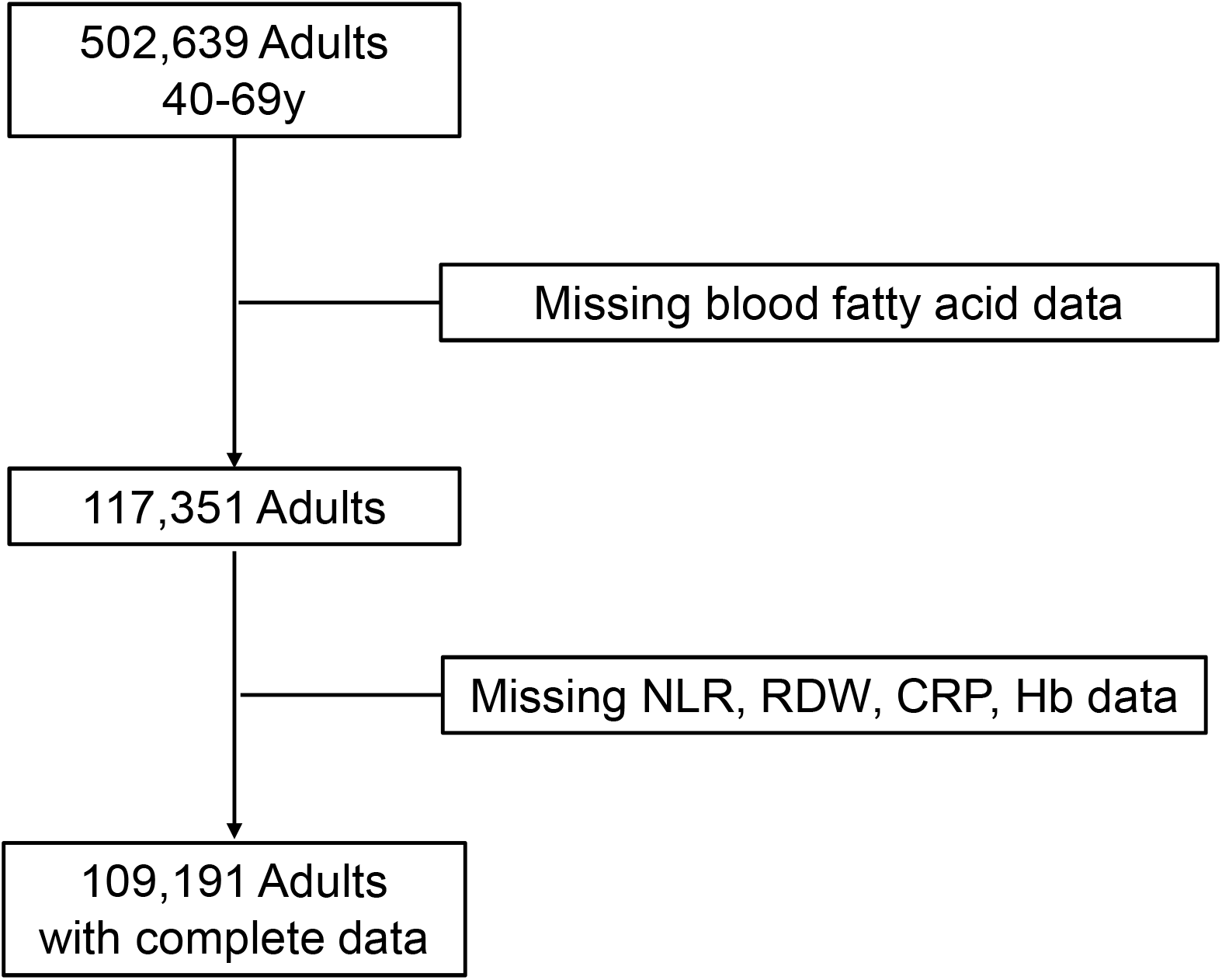
Analytical sample flow chart. Hb, hemoglobin; NLR, neutrophil-lymphocyte ratio; RDW, red blood cell distribution width; CRP, C-reactive protein.

The UK Biobank study was conducted according to the guidelines laid down in the Declaration of Helsinki. The UK Biobank protocol is available online (http://www.ukbiobank.ac.uk/wp114content/uploads/2011/11/UK-Biobank-Protocol.pdf). The University of South Dakota Institutional Review Board reviewed and approved the use of de-identified data for research purposes (IRB-21-147).

### Statistical methods

Sample characteristics are summarized using standard statistical methods (e.g., means, SDs, correlations) with t-tests. Unadjusted linear models (Model 1) were used to predict RDW and NLR values by cubic splines of Omega3% and DHA%. Model 2 adjusted for sex, age, BMI, CRP and Hb values for RDW and for sex, age, BMI, and CRP for NLR. Pearson correlations were used to assess the strength and direction of linear association between covariates and RDW or NLR. Statistical significance was set to 0.05 for all analyses and 95% confidence bands are provided where appropriate.

## Results

Dietary determinants of Omega3% status have been published for the UK Biobank cohort [27]. Thirty-one percent of participants reported using fish oil supplements, and fatty fish were consumed by 56% of the cohort, factors contributing to a mean Omega3% of 4.42% (**Table 3**) and O3I = 5.59% [27]. In this cohort of 109,191 individuals, the correlation between Omega3% and DHA% was r=0.88. Because Omega3% and DHA% were so highly correlated, only the Omega3% results for Model 1 are presented in the main text, with results for DHA% located in **Supplemental Figure 1**. Model 2 results for RDW- and NLR-relationships with Omega3% and DHA% are also located in **Supplemental Figures 2** and **3**, respectively.

**Table 3.**
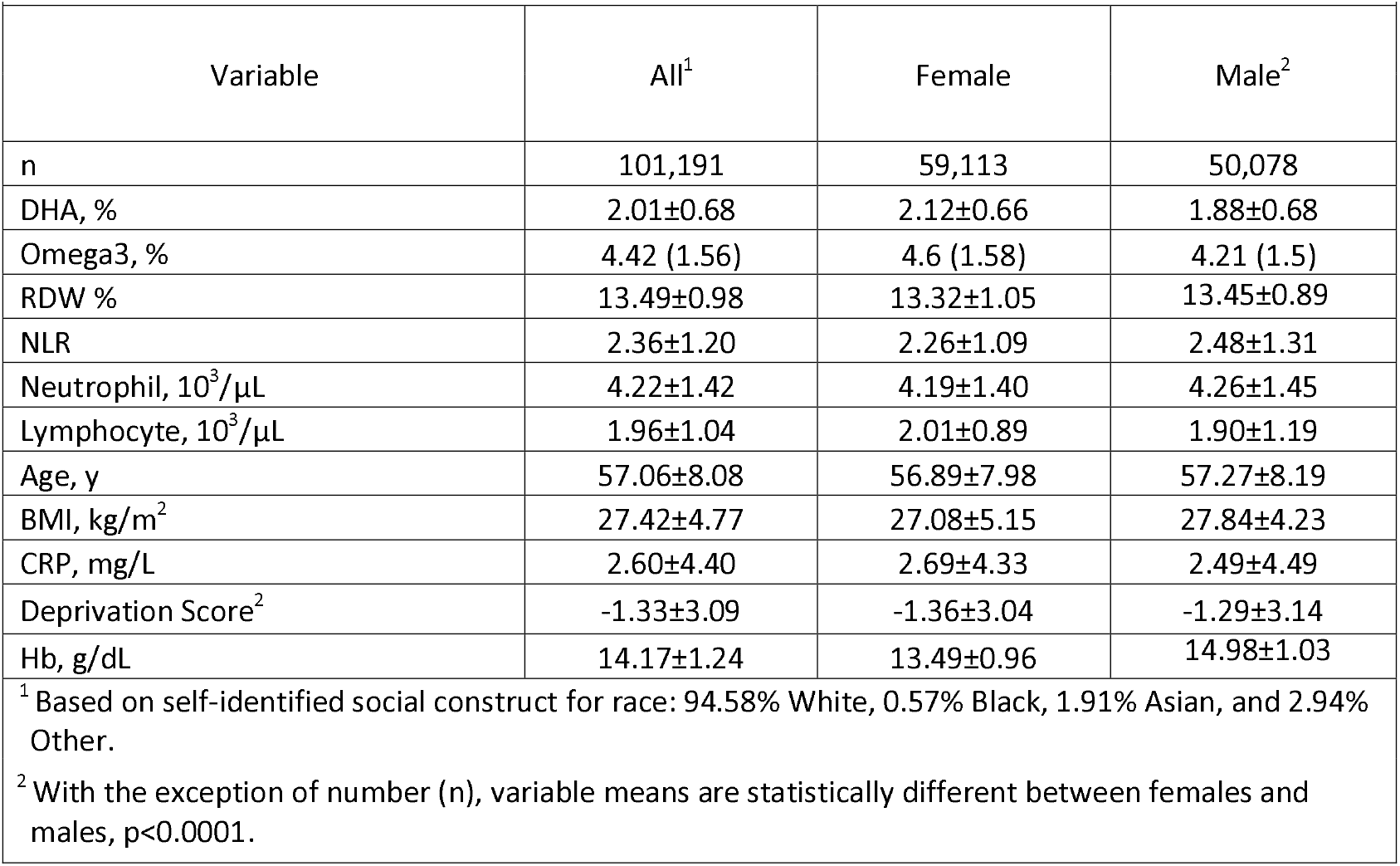
Variable distributions (Mean±SD) for entire UK Biobank cohort.

RDW was significantly (p<0.0001) and inversely associated with Omega3% (**Figure 2A**). Adjustments for age, sex, BMI, CRP and Hb (Model 2) did not significantly change the R^2^ nor the shape of the relationship (**Supplemental Figure 2)**. Excluding 3,698 individuals with anemia (mean cell volume >100fL and/or Hb <13g/dL for males and Hb <12g/dL for females) as done by [22], did not significantly change the R^2^ for the RDW-Omega3% (data not presented).

**Figure 2.**
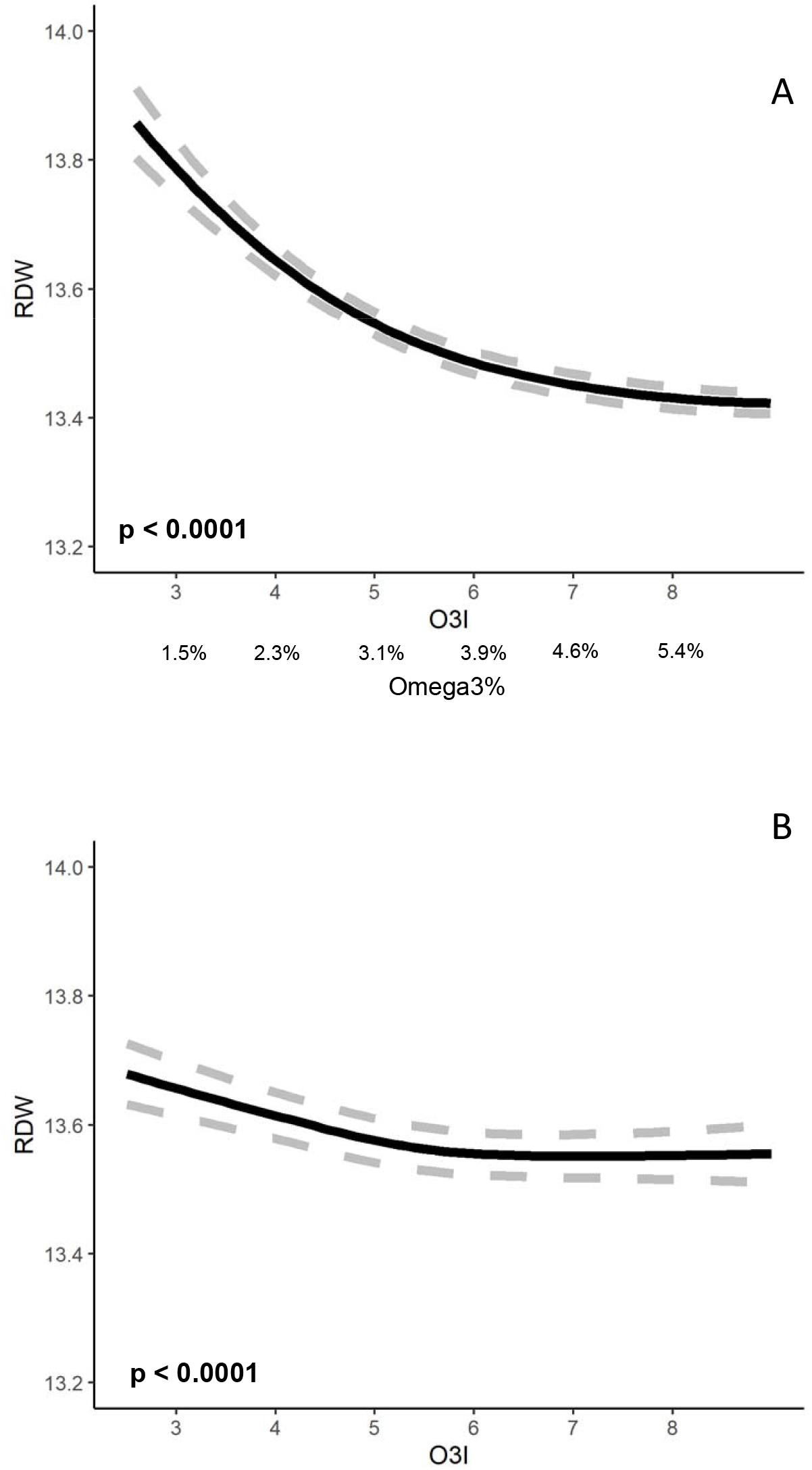
A. Unadjusted red blood cell distribution width (RDW) relationship with omega-3 percentage (Omega3%). Omega-3 Index (O3I) values calculated using the conversion equation [O3I = 1.2791*Omega3%+ 1.0589 (R2=0.59; r=0.77)]. B. Unadjusted RDW relationship with the Omega-3 Index (O3I) adapted from McBurney et al (22).

NLR was significantly (p<0.0001) and inversely associated with Omega3% (**Figure 3A**). Adjustments for age, sex, BMI and CRP (Model 2) did not significantly change the R^2^ nor the shape of the relationship (**Supplemental Figure 2**). Excluding 24,559 individuals with evidence of inflammation, i.e., CRP >3 mg/L, as done by [23], did not significantly change NLR-Omega3% (data not presented).

**Figure 3.**
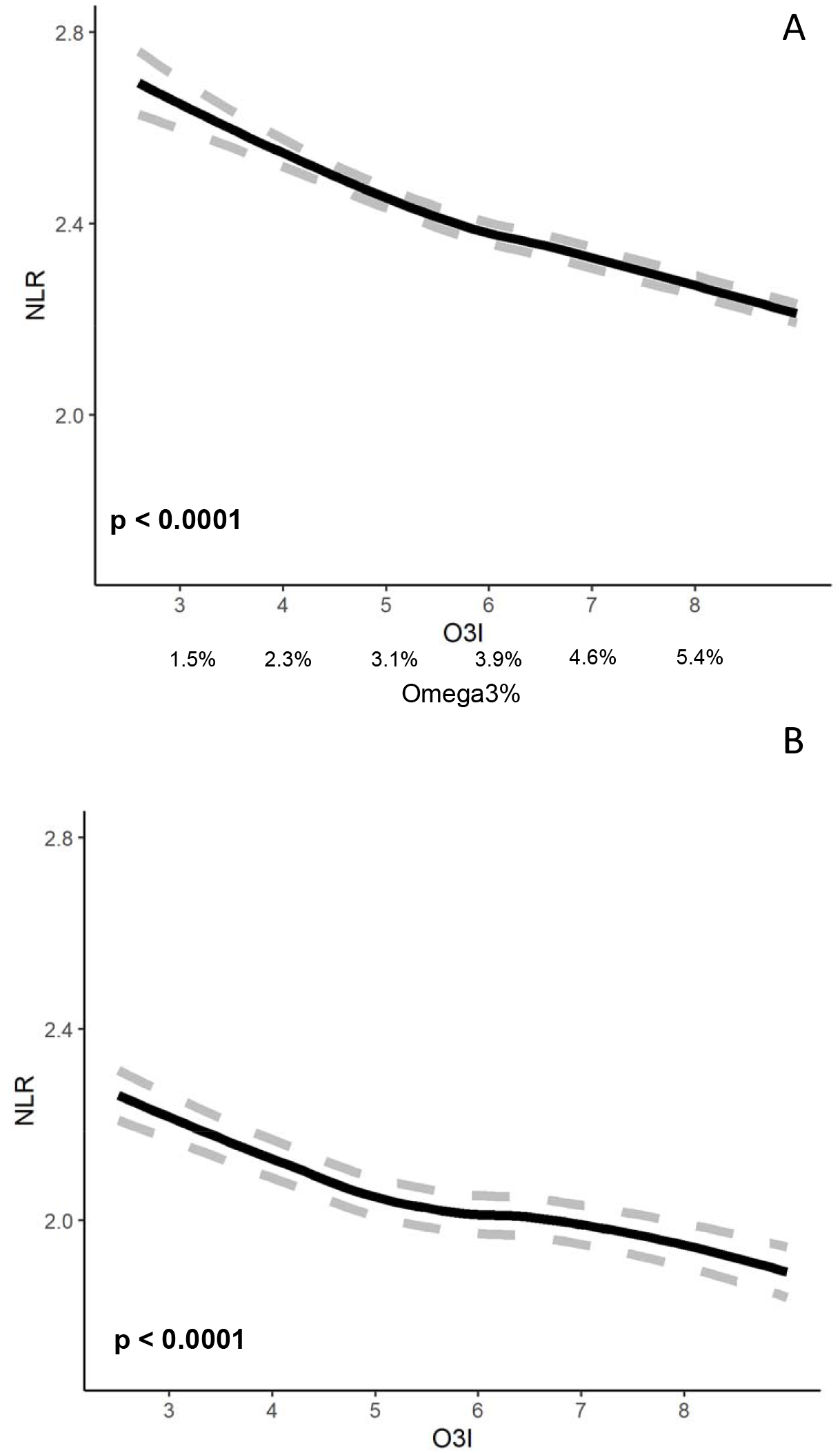
A. Unadjusted neutrophil-lymphocyte ratio (NLR) relationship with omega-3 percentage (Omega3%). Omega-3 Index (O3I) values calculated using the conversion equation [O3I = 1.2791*Omega3%+ 1.0589 (R2=0.59; r=0.77)]. B. Unadjusted NLR relationship with the Omega-3 Index (O3I) adapted from McBurney et al (23).

## Discussion

In this study examining the UK Biobank cohort, we confirm previously reported inverse associations of long chain omega-3 fatty acid concentrations with RDW [22] (**Figure 2**) and NLR [23] (**Figure 3**). The average estimated O3I of 5.59% in the UK Biobank cohort [27] is similar to that (∼5.0%) reported in healthy individuals globally [28,29]. The average RDW in the UK Biobank cohort (13.5%) is similar to values (12.6-12.9%) reported in adults living in the US [30], including those free of cardiovascular disease (NHANES 1994-2004) [31] and without clinical evidence of anemia or inflammation [22]. The UK Biobank average NLR (2.36±1.20, **Table 3**) is similar to the ∼2.2 of the US population [3,6] and a healthy cohort without evidence of anemia or inflammation [23].

Elevated RDW levels predict risk for several adverse health outcomes including death (**Table 1**). In four studies of apparently healthy, free-living individuals, the highest RDW category (Cn; top 20-25% of individuals) has an average HR=1.17 (vs C1) (**Table 4**). Thus, the observed distribution of mean RDW values across values of the Omega3% (13.85-13.40=0.45 units) (**Figure 2A**) corresponds to a HR = 1.06 or a 6% increased risk of all-cause mortality associated with Omega3% when moving from the highest to lowest observed Omega3% values.

**Table 4.** Mortality Hazard Ratio (HR) comparing the highest (top 20-25% of individuals) and lowest (bottom 20-25% of individuals) categories in red blood cell distribution width (RDW) in apparently healthy, free-living individuals.

|  |  |  |  |  |  |  |  |
| --- | --- | --- | --- | --- | --- | --- | --- |
| Outcome | Population | Study | n | RDW (%) Cn-C1<sup>1</sup> | HR (95% CI) | p-value | Citation |
| Coronary heart disease (CHD) deaths, men | Adults ≥20y without diabetes or previous CV | NHANES 1988-1994, USA | 7,201 | 1.10 | 1.19 (1.16-1.22) | <0.001 | [36] |
| CHD deaths, women | Adults ≥20y without diabetes or previous CV | NHANES 1988-1994, USA | 8,259 | 1.10 | 1.17 (1.13, 1.22) | <0.001 | [36] |
| All-cause mortality | Adults ≥40y | NHANES 1988-1994, USA | 15,852 | 1.45 | 1.18 (1.14, 1.22) | <0.001 | [38] |
| All-cause mortality | Adults ≥45y | NHANES 1988-1994, USA | 8,175 | 1.50 | 1.14 (1.10, 1.19) | <0.001 | [39] |
| Average |  |  |  | 1.29 | 1.17 |  |  |
| <sup>1</sup> Highest category (Cn) minus lowest C1. |  |  |  |  |  |  |  |

An elevated NLR has been associated with increased risk of cardiovascular events, lower respiratory disease, influenza, cancer, and mortality (**Table 2**). In two studies of free living, apparently healthy individuals, the average HR for mortality in the highest NLR category (Cn; top 20-25% of individuals) vs the lowest (C1) NLR (**Table 5**) was 1.43. As shown in Figure 3, there was a 0.7 difference in the mean NLR (1.7 to 2.4) across the observed Omega3% range (**Figure 3A**), which is equivalent to a HR = 1.24 or a 24% increased risk of all-cause mortality associated with Omega3% when moving from the highest to lowest observed Omega3% values.

**Table 5.** Mortality Hazards Ratio (HR) comparing the highest (top 20-25% of individuals) and lowest categories (bottom 20-25% of individuals) in neutrophil-lymphocyte ratio (NLR).

|  |  |  |  |  |  |  |  |
| --- | --- | --- | --- | --- | --- | --- | --- |
| Outcome | Population | Study | n | NLR<br>Cn-C1<sup>1</sup> | HR<br>(95% CI) | p-value | Citation |
| All-cause mortality | Adults ≥30y | NHANES 1999-2014, USA | 32,454 | 1.4 | 1.30<br>(1.21, 1.41) | <0.0001 | [6] |
| All-cause mortality | Adults ≥45y, community dwelling | Rotterdam Study, The Netherlands | 8,715 | 1.11 | 1.56<br>(1.39, 1.75) | <0.001 | [44] |
| Average |  |  |  | 1.26 | 1.43 |  |  |
| <sup>1</sup> Highest category (Cn) minus lowest C1. |  |  |  |  |  |  |  |

The primary strengths of this study are the large sample of free-living individuals and that blood samples for RDW, NLR, and Omega3% measurements were taken at the same time. Consistent with previous studies (**Figures 2B** and **3A**) [22,23], monotonic inverse relationships were observed with long-chain omega-3 concentrations (**Figures 2A** and **3A**). The higher RDW and NLR values observed in the UK Biobank cohort may reflect a higher prevalence of inflammation (UKBB mean CRP=2.60±4.40 mg/L vs Health Diagnostic Laboratory mean CRP=1.23± 0.77 mg/L) and differences in age (UKBB mean age =57.1±8.08 y vs Health Diagnostic Laboratory mean = 54.3±14.8 y). Among the limitations are the unavailability of data on levels of other n-3 fatty acids (α-linolenic acid, EPA, and n-3 docosapentaenoic acid) and the potential for other unmeasured (or unconsidered) covariates that might have contributed to the observed associations. The major weakness of this and previous studies [22,23] is their cross-sectional natures which do not provide evidence of causality.

In conclusion, significant inverse relationships with RDW- and NLR-were identified with Omega3% and DHA%. These observations in the UK Biobank cohort using a different analytical method, i.e. NMR, confirm previously reported RDW- and NLR relationships with O3I, i.e. EPA+DHA [22,23]. We propose that low Omega3% may indicate a state that is less resilient and/or more predisposed to disease. We cautiously suggest that RDW- and NLR-relationships with Omega3% are clinically relevant and highly recommend that randomized controlled intervention trials using EPA and/or DHA be conducted to determine their effects on RDW and/or NLR values, particularly in individuals with less-than optimal long chain omega 3 values at baseline.

## Data Availability

The UK Biobank study is available online.

http://www.ukbiobank.ac.uk/wp114content/uploads/2011/11/UK-Biobank-Protocol.pdf

## Abbreviations

BMI: body mass index
CRP: high-sensitivity C-reactive protein
DHA: docosahexaenoic acid
eO3I: estimated omega-3 index
EPA: eicosapentaenoic acid
Hb: hemoglobin
Omega3%: total omega-3 fatty acid
NLR: neutrophil-lymphocyte ratio
O3I: omega-3 index
NMR: nuclear magnetic resonance
RDW: red blood cell distribution width.

## CRediT authorship contribution statement

**Michael I. McBurney:** Conceptualization, Writing – original draft, Writing – review & editing. **Nathan L. Tintle:** Formal analysis, Writing -reviewing & editing. William **S. Harris:** Conceptualization, Funding acquisition, Writing – review & editing.

