## Supplemental Figures 1-3 for "Lower Omega-3 Status Associated with Higher Erythrocyte Distribution Width and Neutrophil-Lymphocyte Ratio in UK Biobank Cohort"

### Slide 1
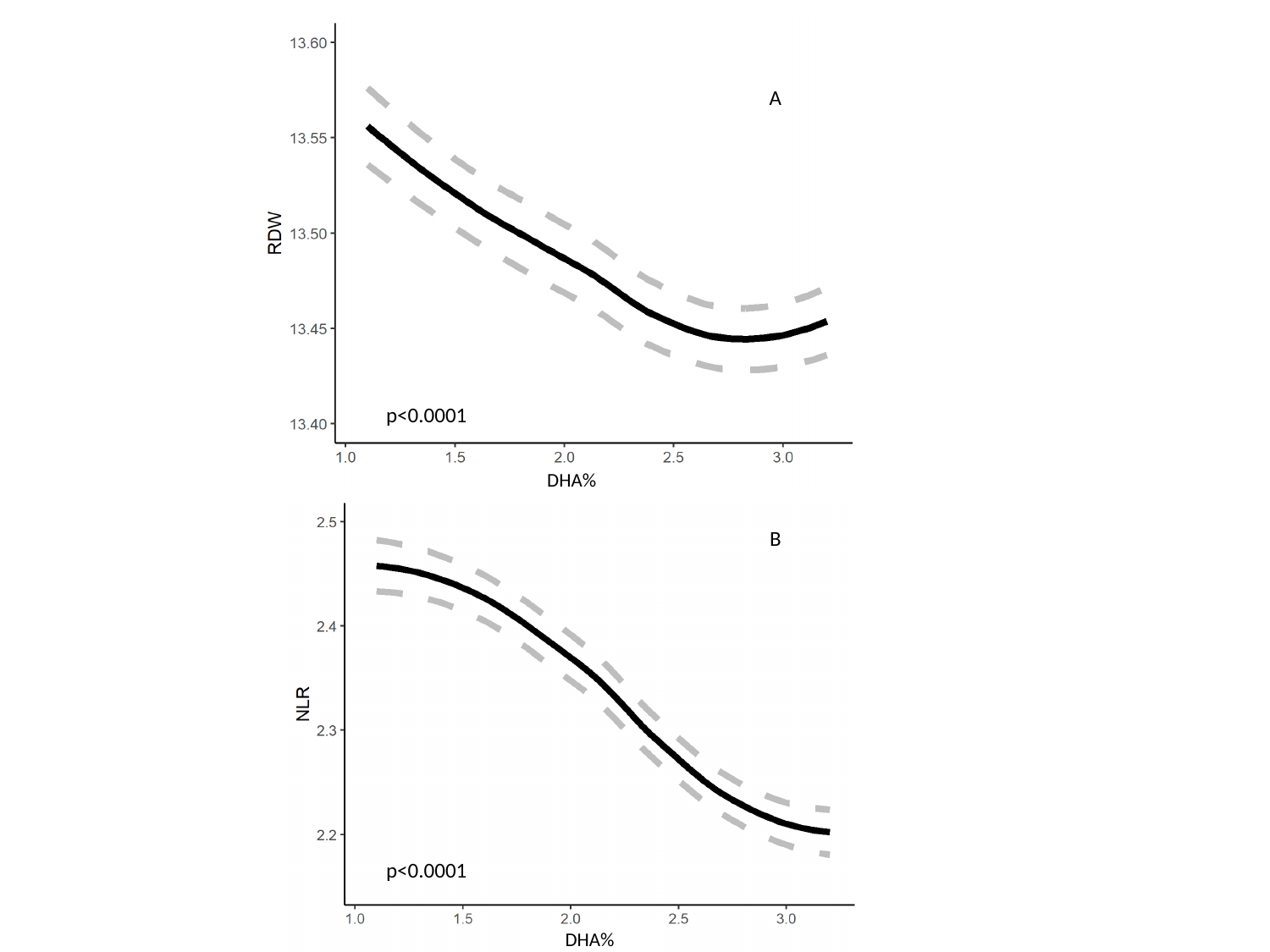

A
p<0.0001
DHA%
B
p<0.0001
DHA%

### Slide 2
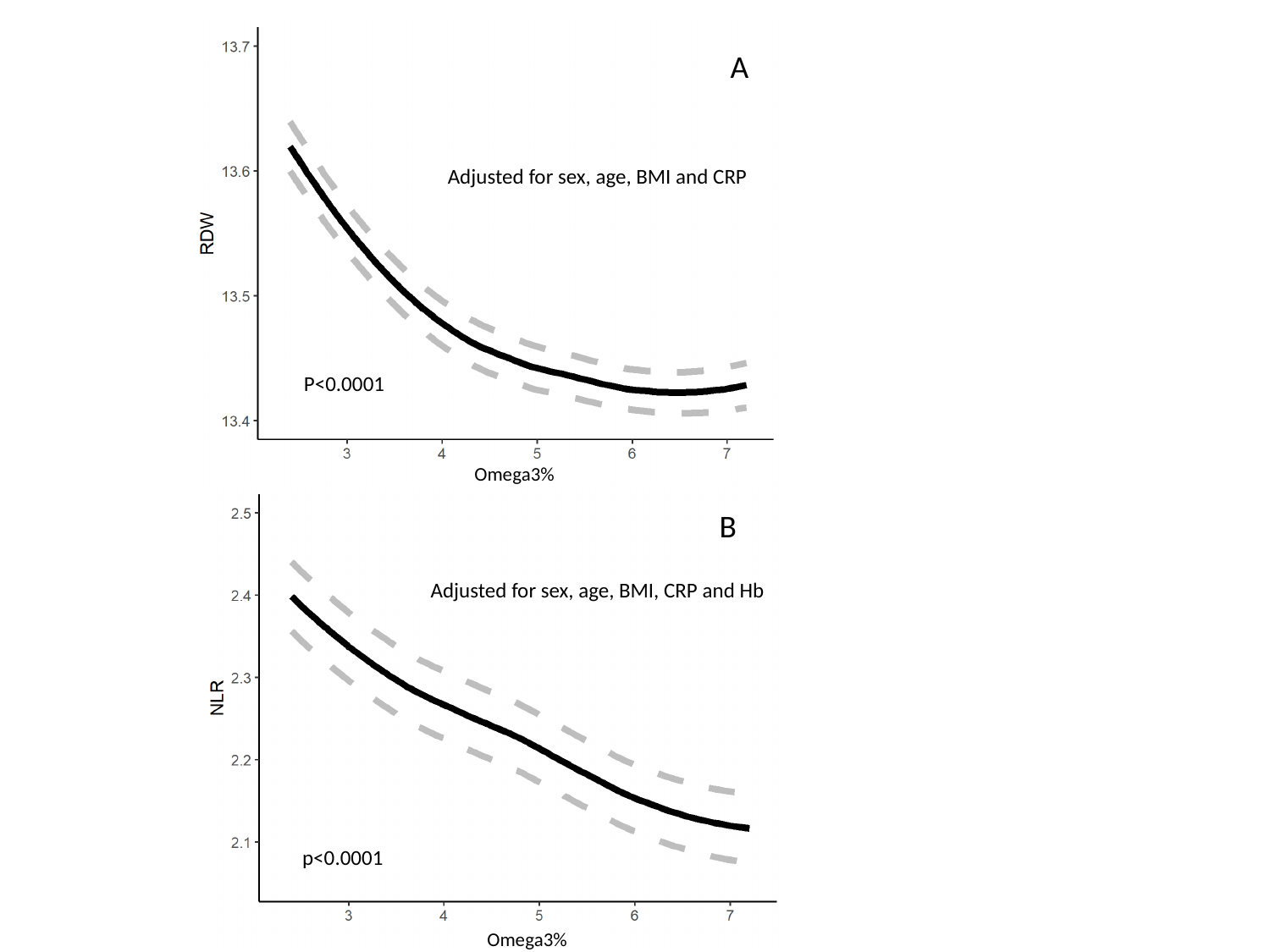

A
Adjusted for sex, age, BMI and CRP
P<0.0001
Omega3%
B
Adjusted for sex, age, BMI, CRP and Hb
p<0.0001
Omega3%

### Slide 3
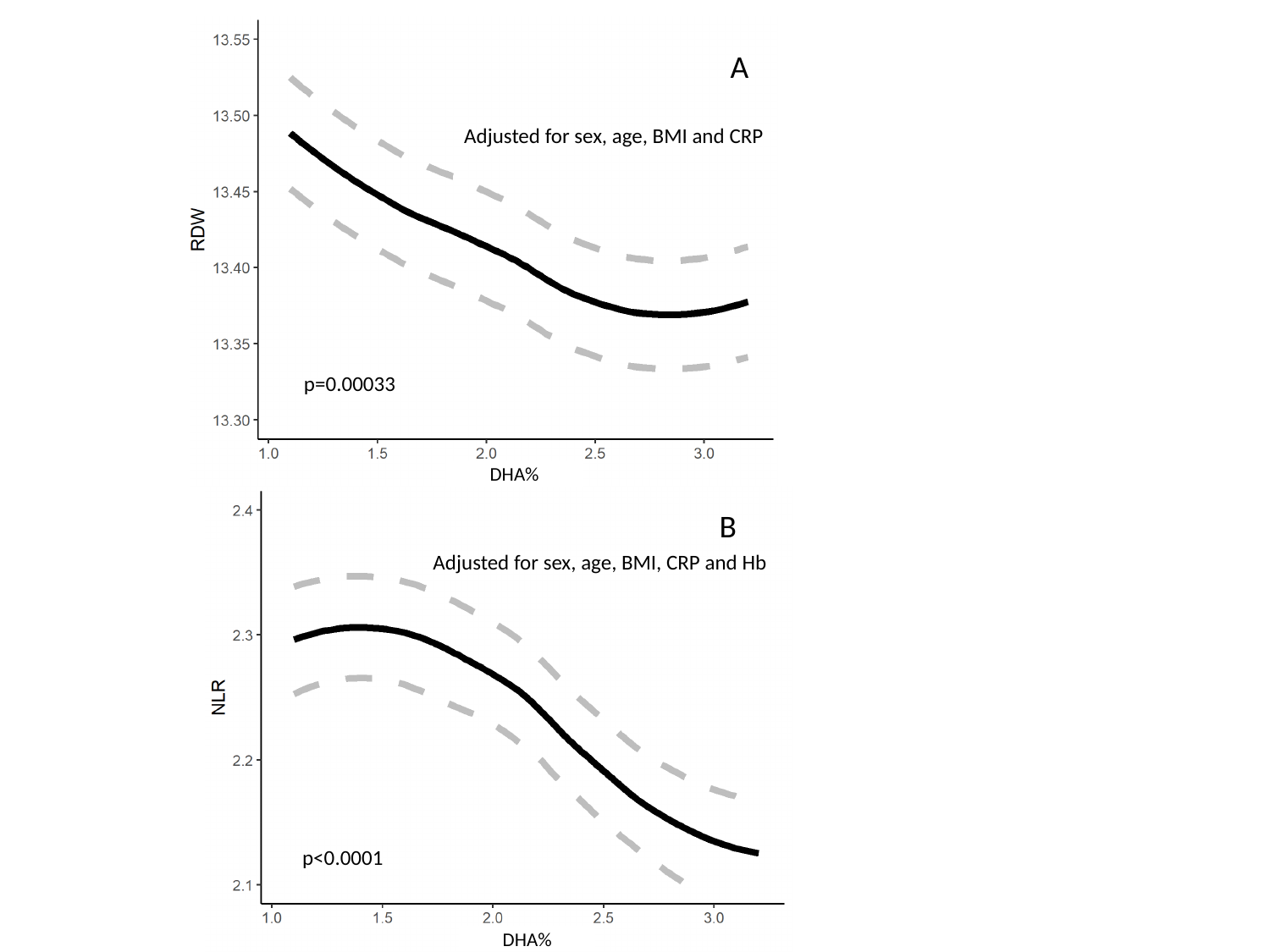

A
Adjusted for sex, age, BMI and CRP
p=0.00033
DHA%
B
Adjusted for sex, age, BMI, CRP and Hb
p<0.0001
DHA%
